# Effectiveness of a mobile health app on initiated antihypertensive medications in patients with untreated hypertension

**DOI:** 10.1101/2023.08.03.23293628

**Authors:** Koichiro Matsumura, Atsushi Nakagomi, Eijiro Yagi, Nobuhiro Yamada, Yohei Funauchi, Kazuyoshi Kakehi, Ayano Yoshida, Takayuki Kawamura, Masafumi Ueno, Gaku Nakazawa, Takahiro Tabuchi

## Abstract

**Background:** Few effective tools have been identified that facilitate the initiation of antihypertensive medications in patients with untreated hypertension. To determine whether a mobile health (mHealth) application facilitates the initiation of antihypertensive medications in patients with untreated hypertension.

**Methods:** We analyzed a large, longitudinal, integrated database mainly comprising middle-aged, working people and their families. The database contained health checkup data, health insurance claims data, and mHealth app data. The mHealth app, kencom, is used to manage daily life logs (i.e. weight, number of steps) and to provide health information tailored to customers. Patients with untreated hypertension were defined using the baseline health checkup data. A multivariable logistic regression analysis was performed to examine the association between use of the mHealth app and the initiation of antihypertensive medications.

**Results:** Among 50 803 eligible patients (mean age, 49 years; 78 % male) with a median follow-up period of 3.0 years. The rate of initiation of antihypertensive medication was 23.4 % vs. 18.5 % (p < 0.0001), which was significantly higher in the mHealth application group (n = 14 879) than in the non-user group (n = 35 924). Multivariable analysis revealed that usage of the mHealth app was associated with initiated antihypertensive medications (odds ratio 1.43, 95 % confidence interval 1.36–1.50).

**Conclusion:** In patients with untreated hypertension, the use of the mHealth app, which was not dedicated to hypertension treatment, was associated with the initiation of antihypertensive medications.

## Introduction

Hypertension is a major risk factor for the development of cardiovascular and renal diseases, causing 8.5 million deaths annually worldwide due to hypertension.^1,2^ Hypertension can be easily detected at community healthcare and primary care facilities and managed using inexpensive drug therapy.^3^ Adequate therapeutic interventions using antihypertensive medications can prevent the development of cardiovascular disease.^4,5^ The number of patients with hypertension worldwide has doubled from 650 million in 1990 to 1.28 billion in 2019, with more than 700 million estimated individuals with untreated hypertension.^6^ To prevent the development of cardiovascular diseases, it is a growing priority worldwide to detect a wide range of patients with untreated hypertension and practice adequate blood pressure control with drug therapy.^7^ Cooperative efforts, such as raising awareness of hypertension in the community and improving access to affordable medical care, are needed to initiate therapeutic interventions for patients.^8^ However, there is little evidence to support the initiation of drug treatment in patients with untreated hypertension.^9,10^

Mobile health apps (mHealth apps) are software applications designed for smartphones and other mobile devices that focus on promoting health and wellness. Leveraging mobile device capabilities, such as sensors, connectivity, and user interfaces, provides a variety of health-related services and support. mHealth apps support not only disease prevention, management, and treatment, but also psychological support and decision-making of patients.^11^ In patients with hypertension, mHealth apps are known to facilitate important recommended lifestyle modifications, reduce the inertia of antihypertensive treatment, and improve patient adherence to medication.^11^ As such, this study aimed to investigate whether an mHealth app is effective in initiating drug therapy in patients with untreated hypertension. In this study, the mHealth app used was kencom, which manages daily life logs (i.e., weight and number of steps) and provides health information tailored to customers.

## Methods

### An outline of the mobile health app and its functionality

The mHealth app (kencom) used in this study is a service developed by DeSC Healthcare Corporation (Tokyo, Japan) and is available on iOS and Android platforms. Free use of the app is available to those who reside in Japan, are 19 years of age or older, and are members of a social insurance labor association, which is one of the main providers of universal health insurance in Japan. The affiliated society-managed employment-based health insurance association pays a subscription fee to the DeSC Healthcare Corporation.

Kencom has five core features.^12^ 1. Display of annual health checkup data: users can view their own longitudinal and detailed health data based on government-initiated annual health checkups without manual input. 2. Daily health data monitoring and goal setting: steps are counted by the built-in pedometer on each smartphone, and data can be synchronized with the kencom app with the user’s consent. Users can set daily physical activity goals, and the kencom app provides feedback based on achievements through self-checks. Users can also manually input their weight, blood pressure, and blood glucose levels into the application. 3. Providing health information: health information based on lifestyle and disease risk is provided to improve health literacy. All original content was peer-reviewed by medical doctors and nutrition experts. 4. Promoting exercise through team competition events: kencom regularly promotes exercise campaigns (arukatsu events). Teams of up to ten users compete for the total number of steps taken over the course of a month. Individual step counts were shared among team members, allowing each member to exercise. Elements of game design were utilized in the event. 5. Incentive system: kencom points are awarded for daily login and use of various services, including exercise campaigns. Points were not awarded for daily activities or goal achievement. From kencom, users can obtain gift certificates that can be used in the marketplace.

### Databases

We used an integrated database provided by DeSC Healthcare Corporation (https://desc-hc.co.jp/en). This database consists of three data sources: the Japanese health checkup database, the Japanese health insurance claims database, and the kencom database. These data were integrated, anonymized, and stored at affiliated regional society-managed employment-based health insurance associations. The DeSC Healthcare Corporation integrates data from different social insurance associations into a large longitudinal database for research purposes. The Japanese health check-up database consists of questionnaire results, physical examinations, biomarker measurements, and imaging examinations performed annually for the majority of adults living in Japan. The Japanese Health Claims database records monthly information on patient demographics, International Classification of Diseases and Related Health Problems, 10th Edition (ICD-10) diagnoses, medical procedures, and medications. The kencom database is aggregated primarily based on daily physical activity data and app usage. The anonymization of the data was based on an "opt-out agreement" between the user and the society’s employment-based health insurance association, which notified the user of any data use and allowed the user to suggest that the data be deleted. This study was approved by the institutional review boards of all participating institutions.

### Target population

Among the participants in the DeSC database from January 2016 to September 2021 (n = 864 413), those with untreated hypertension were selected (n = 66 339) based on baseline health checkup data. Patients with untreated hypertension were defined by the following criteria: those with systolic blood pressure ≥ 140 mmHg or diastolic blood pressure ≥ 90 mmHg and not taking antihypertensive medications upon the baseline medical checkup.^13^ Of these patients, those younger than 20 years (n = 79), with missing blood laboratory data (n = 418), no follow-up data (n = 5 930), and no follow-up data collected more than one year after baseline (n = 9 109) were excluded from this analysis.

### Exposure variable

The exposure variable was mHealth app use. The final analysis categorized the patients into kencom app downloaders (mHealth app users) and non-app downloaders (non-users) (Figure 1). mHealth app users had baseline health checkup data collected within three months prior to the time they downloaded the app. Non-users had baseline data from the first health checkup conducted in the database.

**Figure 1.**
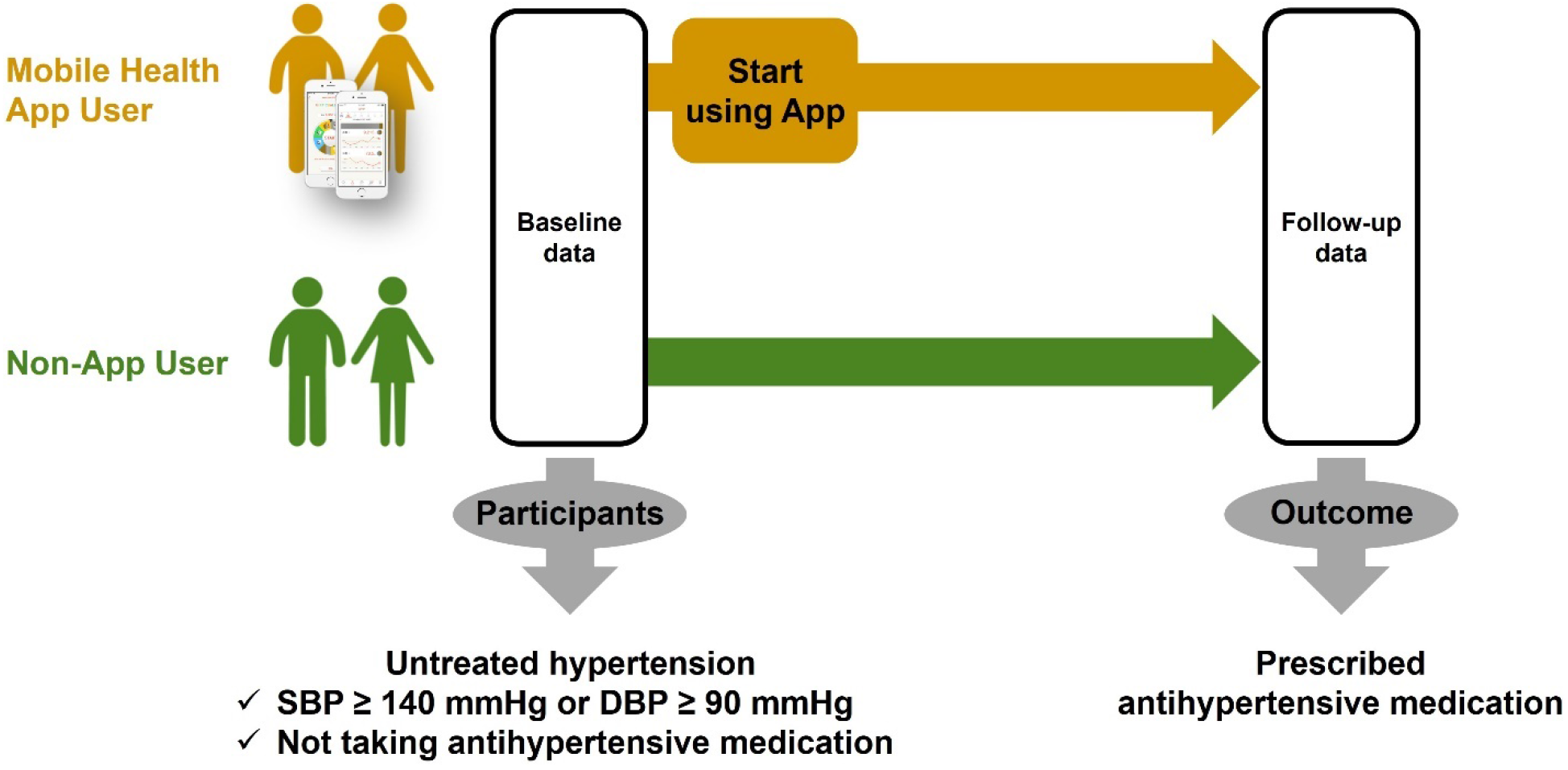
Study flow chart.

### Primary outcome

The primary outcome was the rate of initiation of antihypertensive medication at follow-up. Information regarding the initiation of antihypertensive medication use was collected from the Health Insurance Claims Database.

### Covariates

Age, sex, body mass index, blood pressure, abdominal circumference, blood laboratory data, presence of metabolic syndrome, presence of antihyperglycemic medications, presence of antihyperlipidemic medications, medical history, and life history were obtained from the baseline health checkup data. We used the Japanese criteria for metabolic syndrome.^14^ The criteria were abdominal obesity (abdominal circumference ≥ 85 cm in men and ≥ 90 cm in women) and essential criterion, as well as any two of the following three factors: dyslipidemia (triglycerides ≥ 150 mg/dL and/or high-density lipoprotein cholesterol [HDL-C] level < 40 mg/dL or prescribed antihyperlipidemic medications), blood pressure ≥ 130/85 mmHg or prescribed antihypertensive medication, and fasting glucose ≥ 110 mg/dL. Baseline blood pressure was graded according to 2018 European Society of Cardiology/European Society of Hypertension guidelines; Grade I: SBP 140–159 mmHg or DBP 90–99 mmHg, Grade II: SBP 160–179 mmHg or DBP 100–109 mmHg, and Grade III: SBP ≥ 180 mmHg or DBP ≥ 110 mmHg.^15^ Following this, four lifestyle questionnaires were collected from baseline and follow-up health checkup data: light sweaty exercise ≥ 1 year (do you exercise at least twice a week and for at least 1 year with light sweaty activity for at least 30 minutes per session?), walking or equivalent physical activity ≥ 1 hour/day (do you take walking or equivalent physical activity in your daily life for at least 1 hour/day?), eating dinner within 2 hours before bedtime (do you eat dinner within 2 hours before bedtime at least three times a week?), and improved lifestyle habits such as exercise and diet (Are you intending to improve your lifestyle habits such as exercise and eating habits?). The responses to “improve lifestyle habits such as exercise and diet” were as follows: no intention to improve (precontemplation), will improve within 6 months (contemplation), will improve within 1 month and have started gradually (preparation), have been working on improvement for 6 months or less (action), and have been working on improvement for more than 6 months (maintenance).

### Statistical analysis

In the descriptive statistics, means ± standard deviations were used for continuous variables, and numerical values and percentages were used for categorical variables. We used multivariate logistic regression analysis to examine the association with the use of initiated antihypertensive medications. After estimating the logistic regression models, we calculated the average marginal effects (AMEs), which estimated the average of the predicted changes in the probability of initiating antihypertensive medication, controlling for other covariates. Four sensitivity analyses were performed to examine the heterogeneity of associations. First, a subgroup analysis was conducted to evaluate the adjusted odds ratios for all covariates. This analysis was conducted using formal assessments for effect-measure modification on multiplicative scales (coefficients of product terms in logistic models). Second, the rate of initiation of antihypertensive medications between mHealth app users and non-users was assessed and stratified by hypertension classification. Third, mHealth app users were more likely than non-users to be aware of the need to improve their lifestyle at baseline. Accordingly, responses to the "improve lifestyle habits such as exercise and diet" question collected at baseline were stratified into three groups: those responding to action or maintenance, those responding to preparation, and those responding to precontemplation or contemplation. Finally, the kencom application regularly promoted exercise campaigns (Arukatsu event), and the mHealth use group was stratified by whether they participated in exercise campaigns (Arukatsu event).

In this study, there were variables with missing values. To address the potential bias resulting from missing data, we conducted multiple imputations using the Markov Chain Monte Carlo method under the assumption that data were missing at random conditions associated with all variables in Supplementary Table 1.^16^ After generating 100 imputed datasets using all variables, we performed the analyses described above and combined the effect estimates using Rubin’s rule.^17^ All statistical analyses were performed using the JMP version 16.0 (SAS Institute) or STATA 17.0 software (STATA Corp. LLC Collage Station, TX, USA). A two-tailed alpha value of 0.05 was considered statistically significant.

## Results

### Patient characteristics

In total, 50 803 patients were included in the final cohort. The median observation period was 3.0 years. Patient backgrounds are shown in Table 1. The mean patient age was 49 years and 78 % of the patients were male. Metabolic syndrome was present in four patients. The answers to the question "improve lifestyle habits such as exercise and diet" are shown in the stage of health behavior changes. The proportion of patients who answered "action" and "maintenance" who were already implementing health behaviors to improve their lifestyle was approximately one-fourth of the overall patients, and was higher among mHealth app users compared to non-users (24.5 % vs. 28.1 %).

**Table 1.** Baseline characteristics.

|  | n | Total cohort | mHealth app |  |
| --- | --- | --- | --- | --- |
|  |  |  | User | Non-user |
| Age (years) | 50 803 | 49 ± 10 | 48 ± 8 | 50 ± 10 |
| Men | 50 803 | 39 412 (77.6) | 12 982 (87.3) | 26 430 (73.6) |
| BMI ≥ 25.0 kg/m<sup>2</sup> | 50 801 | 20 751 (40.8) | 6 398 (43) | 14 353 (40) |
| Waist circumference (cm) | 49 751 | 86 ± 10 | 86 ± 10 | 86 ± 10 |
| SBP (mmHg) | 50 803 | 144 ± 11 | 143 ± 11 | 144 ± 11 |
| DBP (mmHg) | 50 803 | 92 ± 8 | 93 ± 8 | 91 ± 9 |
| Hypertension classification | 50 803 |  |  |  |
| Grade I |  | 42 680 (84) | 12 459 (83.7) | 30 221 (84.1) |
| Grade II |  | 6 755 (13.3) | 2 045 (13.7) | 4 710 (13.1) |
| Grade III |  | 1 368 (2.7) | 375 (2.5) | 993 (2.8) |
| Laboratory data |  |  |  |  |
| Triglyceride (mg/dL) | 50 803 | 136 ± 111 | 138 ± 111 | 135 ± 112 |
| HDL cholesterol (mg/dL) | 50 802 | 61 ± 17 | 60 ± 16 | 61 ± 17 |
| Fasting blood glucose (mg/dL) | 43 425 | 99 ± 19 | 99 ± 18 | 100 ± 19 |
| Metabolic syndrome | 43 425 | 10 634 (24.5) | 3 441 (25.4) | 7 193 (24.1) |
| Receiving antihyperglycemic medications | 50 802 | 1 494 (2.9) | 375 (2.5) | 1 119 (3.1) |
| Receiving antihyperlipidemic medications | 50 801 | 3 492 (6.9) | 920 (6.2) | 2,572 (7.2) |
| History of cardiovascular disease | 42 511 | 678 (1.6) | 188 (1.4) | 490 (1.7) |
| History of stroke | 42 508 | 277 (0.7) | 77 (0.6) | 200 (0.7) |
| Current smoking | 50 793 | 13 677 (26.9) | 3 638 (24.5) | 10 039 (28) |
| Alcohol consumption | 44 157 | 14 302 (32.4) | 4 863 (36.3) | 9 439 (30.7) |
| Stage of health behavior changes | 41 925 |  |  |  |
| Precontemplation |  | 9 474 (22.6) | 2 461 (19.2) | 7 013 (24.1) |
| Contemplation |  | 15 310 (36.5) | 4 787 (37.4) | 10 523 (36.1) |
| Preparation |  | 6 419 (15.3) | 1 955 (15.3) | 4 464 (15.3) |
| Action |  | 4 381 (10.4) | 1 491 (11.7) | 2 890 (9.9) |
| Maintenance |  | 6 341 (15.1) | 2 098 (16.4) | 4 243 (14.6) |

Data are presented as mean ± SD or n (%). BMI: body mass index.

### Primary endpoint

The follow-up health checkup data and health insurance claims databases were used to assess the rate of antihypertensive medication use (Figure 2). mHealth app users had a significantly higher rate of antihypertensive medication initiation than non-users (23.4 % vs. 18.5 %, p < 0.0001).

**Figure 2.**
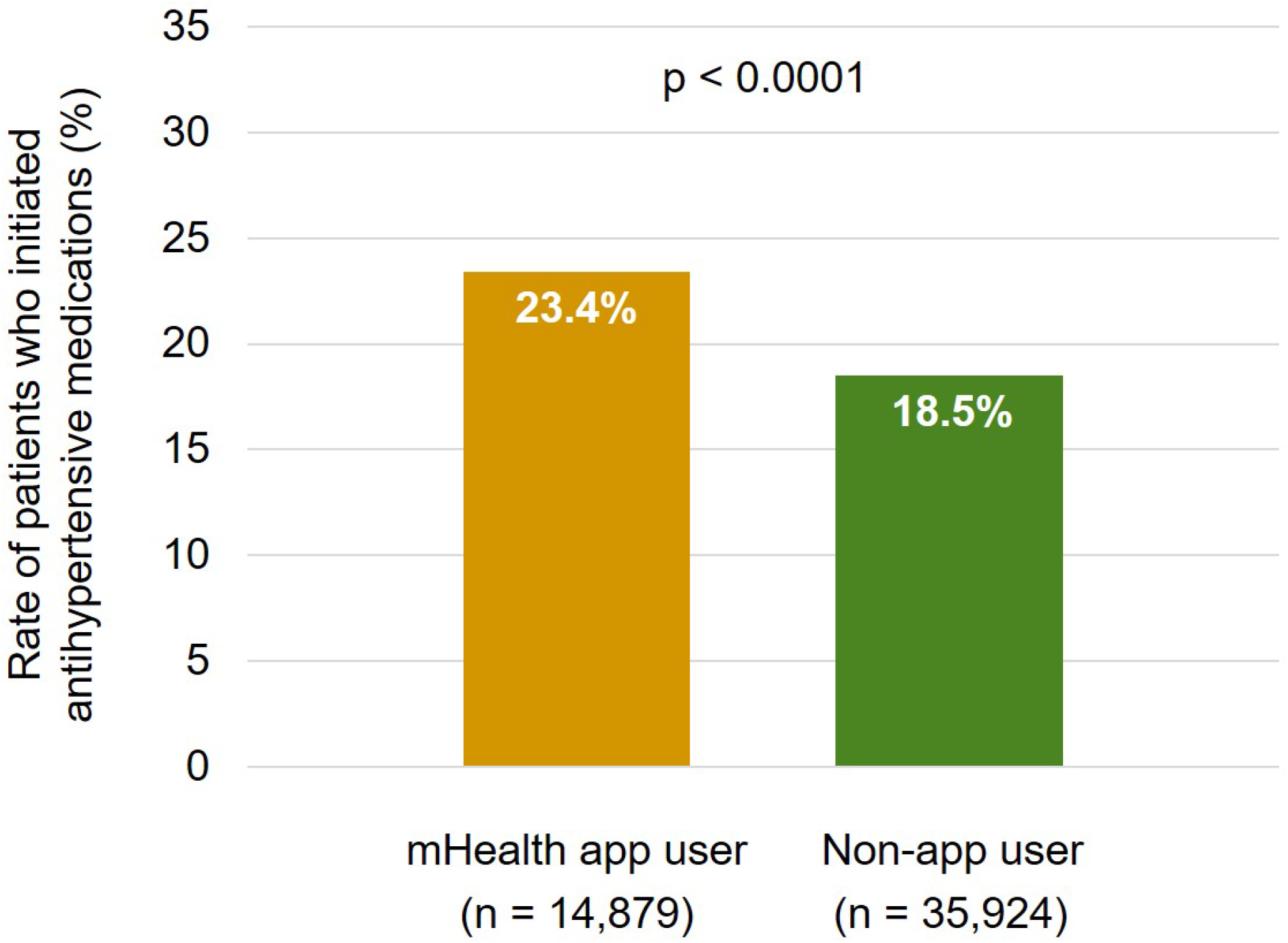
Percentage of patients who initiated antihypertensive medications.

### Multivariable analysis

Multivariate analysis using a logistic regression model showed that mHealth app usage was associated with the initiation of antihypertensive medications (odds ratio, 1.43; 95 % confidence interval, 1.36–1.50; Table 2). The predicted probability of antihypertensive medication initiation was on average 5.3 % greater in users than in non-users. The results of the multivariate analysis for the other variables are presented in Supplemental Table 2.

**Table 2.**
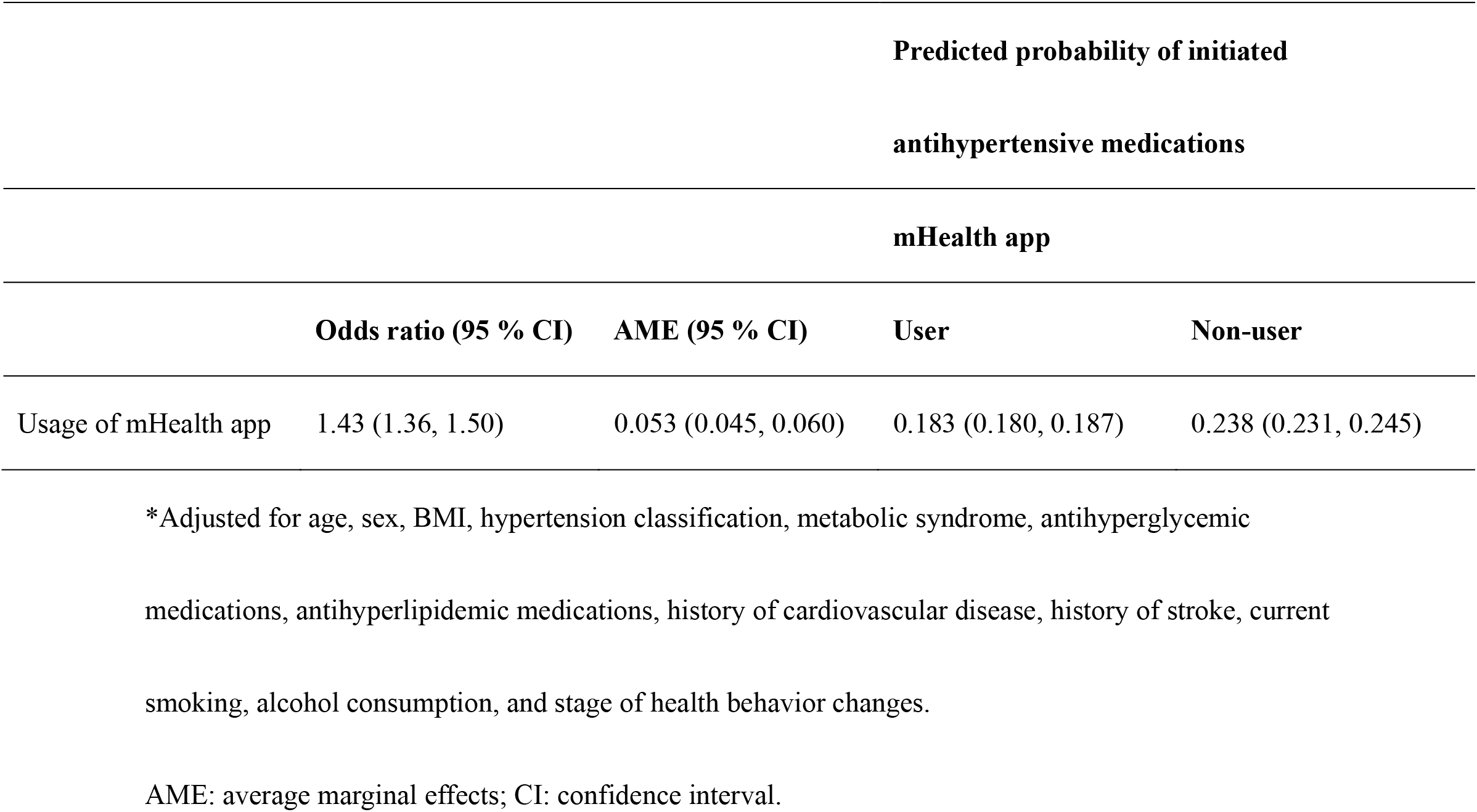
Associations between usage of mHealth app and initiating medication for hypertension*.

### mhealth app and lifestyle modification behaviors

The rate of patients who responded "no" to each of the four lifestyle questionnaires at baseline but responded "yes" at follow-up was examined. The rate of patients who had not performed light sweaty exercise ≥ 1 year who were performing it at follow-up as an mHealth app user: 12.4 % (1 594 of 12 906) vs. non-user: 10.0 % (2 889 of 28 878), p < 0.0001. Likewise, the rate of walking or equivalent physical activity ≥ 1 hour/day in an mHealth app user was 15.7 % (2 042 of 12 993) vs. 12.8 % in non-users (3 754 of 29 319), p < 0.0001. The rate of respondents who did not eat dinner within 2 hours before bedtime was 13.7 % in mHealth app users (1 850 of 13 533) vs. 10.3 % in non-users (3 256 of 31 591), p < 0.0001. For improvements in lifestyle habits such as exercise and diet, mHealth app users were 18.9 % (2 397 of 12 685) vs. 14.4 % of non-users (4 121 of 28 559), p < 0.0001. All questionnaires indicated that mHealth app users demonstrated a significantly higher rate of lifestyle changes at follow-up than non-users.

### Sensitivity analysis

Figure 3 shows the results of subgroup analyses for all covariates. We found that the odds ratios in the younger group were greater than those in the older group. In addition, the odds ratio of patients without metabolic syndrome was higher than that of patients with metabolic syndrome. No effect modifications were observed for the other covariates.

**Figure 3.**
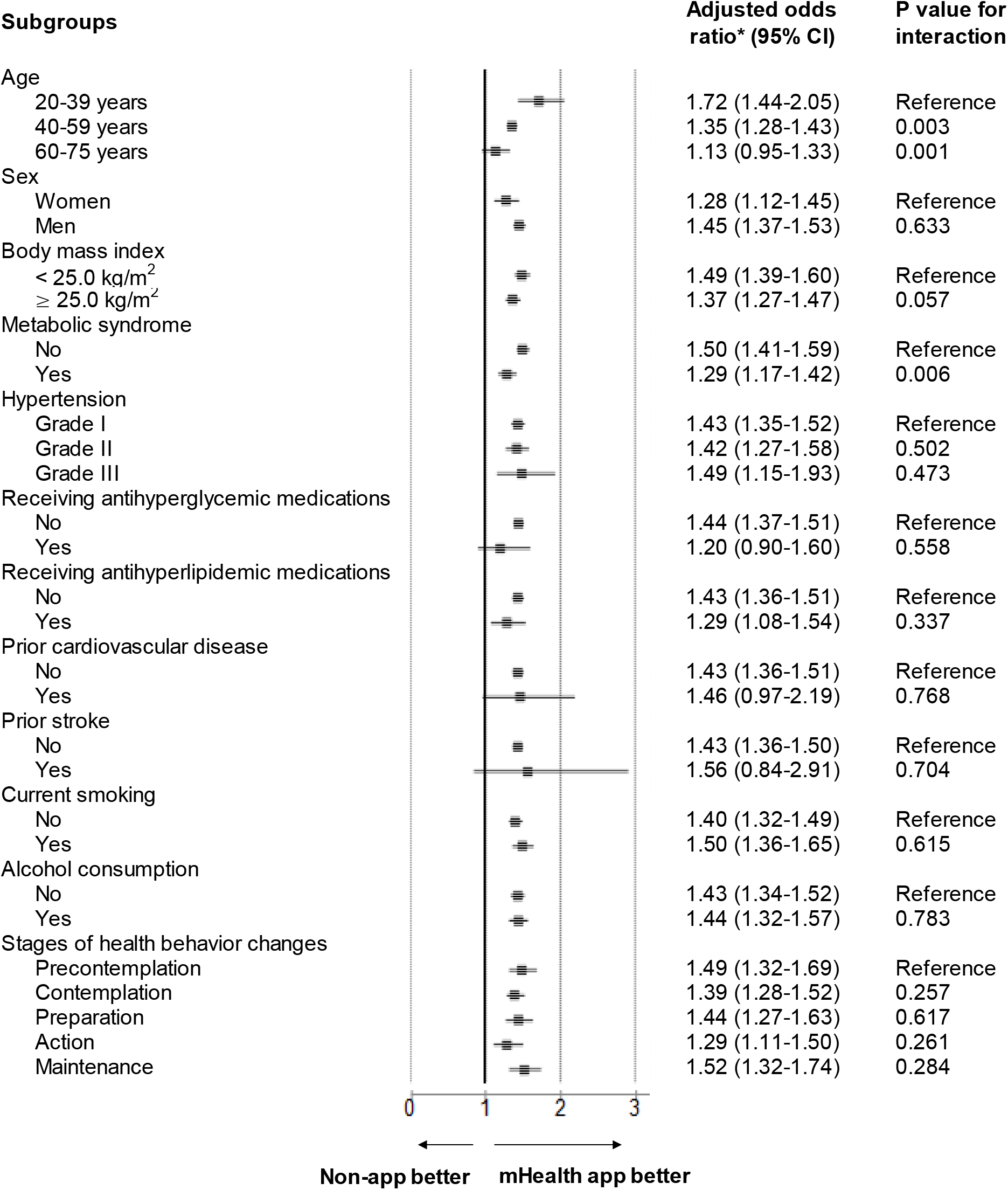
Primary outcome analyzed according to subgroup.

When stratified by hypertension classification, mHealth app users were significantly more likely to initiate antihypertensive medication use regardless of severity (mHealth app users vs. non-users: Grade I, 17.3 % [1 885 of 10 890] vs. 13.1 % [3 340 of 25 518]; Grade II, 37.0 % [1 337 of 3 610] vs. 29.1 % [2 734 of 9 403]; Grade III, 66.5 % [252 of 379] vs. 56.9 % [571 of 1 003], Supplemental Figure).

Among the mHealth app users, we evaluated the rate of initiation of antihypertensive medications among those who participated in the exercise campaigns (Arukatsu event) and those who did not. The results showed that 23.0 % (1 340 of 5 820) of those promoting exercise campaigns (Arukatsu event) and 23.6 % (2 134 of 9 059) of nonparticipants started antihypertensive medications, with comparable rates in both groups (p = 0.45).

In response to the question "improve lifestyle habits such as exercise and diet" at baseline, the rate of initiated antihypertensive medications was compared between mHealth app users and non-users, then stratified into three groups. Among those who responded "precontemplation" or "contemplation," the rate of initiated antihypertensive medications was significantly higher among mHealth app users (24.1 % [865 of 3 589]) compared with non-users (18.8 %[1 342 of 7 133]) (p < 0.0001). Similarly, in those who responded with “preparation,” the rate of initiated antihypertensive medications were 25.6 % (500 of 1 955) vs. 19.4 % (864 of 4 464) (p < 0.0001). In those who responded with “action” or “maintenance”, 23.5 % (1 700 of 7 248) vs. 18.0 % (3 161 of 17 536) (p < 0.0001), respectively, initiated hypertensive medications. Both were significantly higher among mHealth app users.

## Discussion

In this study, we examined whether an mHealth app (kencom) that is not dedicated to hypertension treatment could help promote the initiation of antihypertensive treatment among patients with untreated hypertension, mainly in middle-aged working people and their families. The mHealth app users had higher rates of initiated antihypertensive medication use than non-users. The use of the mHealth app was associated with the initiation of antihypertensive medication use in the multivariable analysis. In the group of patients with low motivation to improve their lifestyle at baseline, a significantly greater number of mHealth app users had improved their lifestyle at follow-up than non-users. The subgroup analysis suggested that the mHealth app may have a greater impact on the initiation of antihypertensive medications in younger patients or in patients without metabolic syndrome. Regardless of baseline blood pressure levels or motivation to make lifestyle changes, mHealth app users consistently initiated antihypertensive medications at higher rates. These results suggest that the mHealth app may have been effective in facilitating the initiation of antihypertensive medications and may have influenced the health modification behaviors of patients with untreated hypertension.

Health literacy refers to the ability to obtain, read, understand, and use medical information to make informed health decisions and plays an important role in the implementation of medication therapy.^18^ People with higher health literacy are more likely to understand the risks associated with hypertension and the importance of regular blood pressure monitoring, healthy lifestyle habits, and medical care.^18^ They are also more likely to understand the benefits and potential side effects of different treatments and make informed decisions regarding their healthcare. In contrast, individuals with low health literacy may not fully understand the risks associated with hypertension, how to measure blood pressure, or how to manage their condition by developing healthy lifestyle habits. Consequently, hypertension may remain untreated, leading to serious health problems.^9^ Improving health literacy includes raising awareness of diseases and their risks, providing clear and concise information about prevention and treatment options, and facilitating access to healthcare services. By improving health literacy, individuals can better understand their health status and take action to prevent and manage hypertension. Additionally, it can lead to improved health and quality of life.

The effectiveness of mHealth app utilization has been reported to improve health literacy and adherence.^19-21^ In our study, mHealth app users improved their exercise and eating habits at a higher rate than non-users, which may result from the ability of mHealth to improve health literacy. This change in patient perception may have facilitated hypertension treatment. There are various interventions to improve adherence to the treatment of chronic diseases, and mHealth includes multiple intervention components such as informational, behavioral, and social interventions.^9^ Methods that included additional intervention components had a greater effect on adherence. For effective interventions, it is important to have continuous and sustained contact with patients and combine strategies tailored to their needs. Therefore, mHealth can be considered an effective intervention tool.

Previous studies have reported that the factors associated with the subgroup not receiving antihypertensive treatment include younger age, better health, non-obese, no history of cardiovascular disease, no diabetes, and no visits to a primary care physician.^10^ These patients are a common subgroup with inadequate awareness of hypertension and poor blood pressure control.^13^ In our study, the mHealth app was effective in initiating antihypertensive medication in these patient populations. Younger patients were less resistant to mHealth apps and were more familiar with their use than older patients, which may explain why mHealth apps were more effective in younger patients. Furthermore, the older population has a higher prevalence of chronic diseases than the younger population, and may visit a hospital and initiate treatment regardless of the mHealth app intervention. It has been suggested that mHealth apps are more effective in patients with no history of chronic disease and a greater distance from their healthcare providers.

### Strengths and limitations

Most existing research on mHealth apps for hypertensive patients is dominated by studies reporting the effect of mHealth app use on blood pressure reduction and medication adherence in patients who have visited a healthcare provider for hypertension.^22^ However, a major issue with untreated hypertension is that patients with hypertension identified in the preliminary stages of visiting a healthcare provider do not tend to visit a hospital for further care thereafter, and remain untreated.^23,24^ Therefore, it is very important that hypertension is detected during health checkups and in the local community before the patient is referred to a medical facility for treatment. This study is of high clinical relevance, as it highlights the effectiveness of mHealth apps for untreated hypertensive patients in the preliminary stage of visiting a medical facility. This cohort study was conducted using a fairly large sample size among studies that investigated the effectiveness of mHealth apps for hypertension treatment.^22^ Furthermore, the analyzed population was mainly middle-aged working people and their families, who could fully benefit from the early initiation of antihypertensive treatment, confirming that the study is of high clinical utility. Finally, the antihypertensive medication status was collected from receipt data and was highly accurate. The results of this study are of great social significance, as they demonstrate the effectiveness of the mHealth app as a trigger for starting antihypertensive medication in patients with untreated hypertension. This study has several limitations. First, our study participants were primarily healthy Japanese individuals; thus, our results may not be generalizable to other populations. Second, we could not completely exclude the possibility of unmeasured confounding factors. However, in the sensitivity analyses, mHealth apps showed good homogeneity, and randomized controlled trials are required to infer the causal effects of mHealth app use on health outcomes. The current study suggests that among patients with untreated hypertension, mainly composed of middle-aged working people and their families, the use of mHealth apps, managing daily life logs (i.e., weight, number of steps), and providing health information tailored to customers may be an effective means of facilitating the initiation of antihypertensive medications. Younger patients or those without metabolic syndrome may receive a greater contribution from the mHealth app. Promoting antihypertensive treatment in the working population could reduce the future development of cardiovascular diseases.

## Data Availability

The datasets generated and analyzed during the current study are not publicly available due to limitations of ethical approval involving patient data and anonymity but are available from the corresponding author upon reasonable request.

## Nonstandard abbreviations and acronyms

mHealth: mobile health
AMEs: average marginal effects

## Acknowledgments

We would like to thank Editage (www.editage.com) for the English language editing.

## Author contributions

All listed authors have made substantial contributions to the conceptualization, data collection/analysis and/or writing of this manuscript.

## Novelty and Relevance

### What Is New?

Identifying patients with untreated hypertension and helping them practice adequate blood pressure control is important to prevent them from developing cardiovascular disease. However, few strategies are effective for initiating antihypertensive medications in patients with untreated hypertension.

### What Is Relevant?

Among patients with untreated hypertension, middle-aged working people and their families who used mHealth app demonstrated higher rates of initiated antihypertensive medications than non-users. Additionally, using mHealth app may significantly impact initiated antihypertensive medications among younger patients or those without metabolic syndrome.

## Clinical/Pathophysiological Implications

The mHealth app may facilitate the initiation of antihypertensive medications in patients with untreated hypertension.

